# Guangzhou Diabetic Eye Study: rationale, design, methodology and baseline data

**DOI:** 10.1101/2020.02.18.20024778

**Authors:** Wei Wang, Miao He, Xia Gong, Lanhua Wang, Jie Meng, Yuting Li, Kun Xiong, Wangting Li, Wenyong Huang

## Abstract

**Purpose:** The incidence and risk factors for diabetic retinopathy (DR) in southern China remains unclear. This project aims to explore the onset and progression of DR and their determinants through a prospective cohort in South China.

**Methods:** The Guangzhou Diabetic Eye Study (GDES) recruited patients with type 2 diabetic patients registered in the community health centres in Guangzhou, China. Patients with history of ocular treatment, severe refractive opacity, or other systemic diseases were excluded. Comprehensive examinations were performed including visual acuity, refraction, ocular biometry, fundus imaging, blood and urine tests. The biological samples were collected for further study.

**Results:** A total of 2305 eligible patients were included in the final analysis. In total, 14.58% of the participants had any DR and 4.25% had vision-threatening DR (VTDR), among which 76 (3.30%), 197 (8.55%), 45 (1.95%) and 17 (0.74%) were classified as mild NPDR, moderate NPDR, severe NPDR and PDR, respectively. There were 93 (4.03%) patients of diabetic macular edema (DME). The presence of any DR was independently associated with a longer duration of DM, higher degree of HbA1C, insulin treatment, higher average arterial pressure, higher concentration of serum creatinine, presence of urinary microalbumin, older age, and lower BMI (all P <0.001). For VTDR, 7 factors were significant: older age, a longer duration of DM, higher concentration of HbA1c, use of insulin, lower BMI, higher concentration of serum creatinine, and high albuminuria (all P<0.05). These factors were also independently associated with DME (all P <0.001).

**Conclusions:** The GDES is the first large-scale prospective cohort study of the diabetic population in southern urban China, which will help to identify newer imaging and genetic biomarkers for DR in this population.

## Introduction

As one of the chronic complications, diabetic retinopathy (DR) is the leading cause of blindness in the working-age population in developed countries. Almost 100% of patients with type 1 diabetes and 60% with type 2 diabetes will ultimately develop DR, despite the significant ethnic variations.^1^ The Multi-Ethnic Study of Atherosclerosis (MESA) in the United States reported that DR prevalence was significantly higher in the African population (36.7%) than in Caucasian (24.8%–37.4%) and Chinese (25.7%), and the Singapore Epidemiology Eye Study (SEED) study found that Indians were 41% more likely to have DR than Chinese, and Malays had the lowest risk.^2, 3^ China has experienced an exponential increase in diabetic patients for the past two decades, with the prevalence climbing from 3.21% in 1996 to 11.6% in 2013.^4^

While a wealth of studies have reported on the incidence and risk factors of DR, they are predominantly focused on type 1 diabetic patients in developed countries but rarely on Chinese patients with type 2 diabetes.^5^ Only two prospective community-based studies have been conducted: the Shanghai Beixinjing Community Study and the Shenyang Fengyutan Community Eye Study.^6, 7^ However, the authors collected only two 45-degree digital retinal photographs and lacked other examination data, such as ETDRS-7-field photographs and optical coherence tomography (OCT). Thus, the results cannot fully depict the onset and progression of DR in China and cannot be directly compared with findings in Western studies.

The population in the Pearl River Delta in China is approximately 100 million. However, epidemiology cohort studies with diabetic population in this area are still lacking to date.^8,9^ Guangzhou, one of the biggest cities in China, has embraced rapid wealth accumulation in the past three decades and differs greatly from the other regions in terms of climate, dietary habits and lifestyle. Its residents enjoy good health insurance, and all diabetic patients are required to register in their communities and attend regularly free physical examinations, making the group an optimal sample to study DR and its complications. The Guangzhou Diabetic Eye Study (GDES) included participants from the diabetic population registered in the communities in Yuexiu district in Guangzhou, and it represents the first large-scale cohort study among diabetic patients in southern rural China. In this article, the intention is to introduce the research background, design, methodology and baseline data for the GDES.

## Materials and methods

### Study design and setting

The GDES is an ongoing community-based prospective cohort study that includes type 2 diabetic patients registered in the community health centres in Yuexiu district in Guangzhou. This district is one of the oldest and centre area of Guangzhou, the population is stable. The Zhongshan ophthalmic centre (ZOC), which is affiliated with Sun Yat-sen University in Guangzhou, locates in this district and is the main provider for eye health services for the city. The communities are stable and the community health centres have established long-term collaboration with ZOC and are fully equipped with a well-established registry system for diabetic patients. The participants were originally recruited in November 2017, and the plan was to follow up on them annually for 5 years. All ocular examinations were performed at ZOC. The study adhered to the Declaration of Helsinki, and approval was granted from the Ethical Review Institute of ZOC. Written informed consent was obtained from all the participants before enrolment.

### Specific aims

The main aims of the study are to (1) evaluate the prevalence, incidence and progression rates of DR among diabetic patients; (2) establish a multi-modality ocular imaging database for DR patients; (3) identify risk factors for DR onset and progression, including demographic indicators, clinical indicators and blood and urine assay indicators; (4) establish a gene database for diabetic patients to examine the association between the sequence variations of candidate genes (e.g. the VEGF, HIF, FTO and EPO genes) or single nucleotide polymorphisms (SNP) with DR in type 2 diabetic patients in southern China; (5) establish a plasma/serum sample bank for future studies; and (6) establish a risk-prediction system for DR onset and progression in type 2 diabetic patients in China, based on the genetic and environmental factors and imaging markers.

### Sample size and power

The primary outcome of the study is the establishment of the progression rate of DR. Previous studies in developed countries/cities, including the United Kingdom Prospective Diabetes Study, the Blue Mountains Eye Study and the Beixinjing Community Cohort Study, showed that the progression rate of DR for a 5-year follow-up period was 25.90%–46.89%.^6,10,11^ Assuming the 5-year progression rate of DR was 30% and considering an acceptable confidence interval of 95% (Z_1-α/2_ = 1.96) and accuracy of 0.05, we estimated the target sample size was 323 individuals (n = z^2^ * p * (1 - p) / e^2^). According to a cross-sectional study conducted in six provinces in China, the prevalence of DR in diabetic patients was 34.08%, and thus we inflated the sample size to 948.^12^ Estimating an attrition rate up to 20%, the target sample size was finally set at 1185 individuals with type 2 diabetes.

### Recruitment strategies

All the eligible participants were recruited from the registered diabetic patients from November 2017 to December 2019, and all of them were referred by doctors in community health centres through advertisements, flyers or phone call and instructed to receive ocular examinations at ZOC. Participants aged 35–85 years with diagnosed type 2 diabetes and ocular-treatment-naïve eyes were included. The diagnosis of diabetes was confirmed by medical records from endocrinologists, insulin treatments, oral medicine, fasting blood glucose≥7.0 mmol/L for at least two consecutive measurements or postprandial blood glucose ≥11.1 mmol/L. The duration of DM was defined as the time from first diagnosis by an endocrinologist to participation in the study. The exclusion criteria were as follows: (1) any severe systemic disease, such as ischemic heart disease, stroke, cancer and kidney disease, or a history of systemic surgery, such as cardiac bypass, thrombolysis and kidney transplantation; (2) any cognitive disorder, mental disorder or failure to finish the questionnaire and all the examinations; (3) combined glaucoma, age-related macular degeneration, glaucoma, vitreous macular disease (vitreous haemorrhage, retinal detachment), amblyopia and other eye diseases; (4) a history of intraocular surgery, such as retinal laser, intraocular anti-VEGF injection, glaucoma surgery, cataract surgery, laser myopia surgery and vitreoretinal surgery history; (5) a failure of mydriasis due to corneal ulcer, severe refractive media turbidity or shallow anterior chamber and angle closure glaucoma.

### Examination procedures

Enrolment and registration: The objectives and procedures of the study were explained in detail to eligible participants after they arrived at ZOC. Prior to all examinations, written informed consent was obtained from all the participants who asked to join the study. Personal information, including national ID card number, name, gender, contact information, address as well as names and contact information of their immediate family member, was recorded.

Visual acuity tests: Visual acuity and refraction tests were performed by qualified optometrists; the right eye was examined first. Naked visual acuity, presenting visual acuity and best corrected visual acuity (BCVA) were measured with the Early Treatment of Diabetic Retinopathy Study (ETDRS) LogMAR E chart (Precision Vision, Villa Park, Illinois, USA) at a distance of 4 meters. The number of letters correctly identified was recorded. If the number was less than 20, visual acuity was examined at a distance of 1 meter. If he/she failed to identify the largest letter, visual acuity was checked by a hand motion followed by a light perception test. The results were recorded as yes or no.

Ocular biometric measurements: Ocular biometric parameters, including axial length (AL), corneal curvature (CC), lens thickness (LT), anterior chamber depth (ACD) and lens thickness (LT) for two eyes, were obtained by an experienced technician using Lenstar LS900 (HAAG-Streit AG, Koeniz, Switzerland) in dark conditions. If the AL surpassed the measurement range of the Lenstar (32 mm), coherence interferometry (IOL Master; Carl Zeiss Meditec, Oberkochen, Germany) was used.

Intraocular pressure (IOP) measurement: IOP was measured before and 30 minutes and 2 hours after mydriasis using Topcon CT-80A non-contact tonometry (Topcon, Japan) for three consecutive measurements with variations less than 0.5 mmHg. The average value was recorded. When the value was higher than 21 mmHg, IOP was remeasured using Goldmann applanation tonometry.

Pupil dilation: Mydriasis was performed by qualified nurses on both eyes using compound tropicamide eye drops (Santen Pharmaceutical, Osaka, Japan) composed of 0.5% tropicamide plus 0.5% phenylephrine, which was administered twice at 5-minute intervals.^13^ Examinations were performed approximately 10–20 minutes after the application when full dilation of the pupil was confirmed. It was considered full mydriasis when the pupil diameter was dilated larger than 6 mm and the light reflex disappeared.

Refraction: Refraction powers, including sphere degree, cylinder degree and axis, were measured after mydriasis with an auto refractometer (KR-8800; Topcon, Japan) by qualified optometrists. Three measurements were obtained for the right eye first followed by the left eye, and only average values were recorded when the disparity of each measurement was less than 0.50 D for the sphere and cylinder powers and 5 degrees for the axis.

Slit lamp and fundus examinations: A slit lamp bio-microscope (BQ-900, Haag-Streit, Switzerland) was used by professional ophthalmologists to evaluate the anterior segment before mydriasis, and any abnormality (e.g. corneal diseases or iris abnormalities) was recorded. The anterior chamber width and depth were recorded to determine the risk of angle closure during mydriasis. The angles were evaluated from the limbal nose and temporal quadrant with a narrow slit beam of about 45 degrees, then all the participants’ eyes were dilated.^14^ In the event of any uncertainty during angle width assessment, an ophthalmologist with higher seniority performed a second examination. After pupil dilation, ophthalmoscopy was used with a slit lamp bio-microscope for posterior segment assessment and any detected abnormalities of the vitreous body and retina were recorded. All the participants were warned against all symptoms of acute angle closure, and the ACD was remeasured after all the ocular examinations. In addition, phone call follow-ups were conducted to inquire about the presence of any symptoms related to angle closure.

Colour retinal photographs: A digital fundus camera (Canon CX-1, Tokyo, Japan) was used to perform standardised 7-field 45° colour retinal photographs for each eye, including the solid shapes of the fovea and optic disc. Only standardised photographs with the maximal definition were kept. The DR was graded according to the International Clinical Severity Scale of Diabetic Retinopathy and Diabetic Macular Edema (Table 1).^15^ Any DR is defined as presence of mild NPDR, moderate NPDR, severe PDR, PDR, or DME. The vision threatening DR (VTDR) is defined as presence of PDR, clinically significant DME, or both.^16^

**Table 1.**
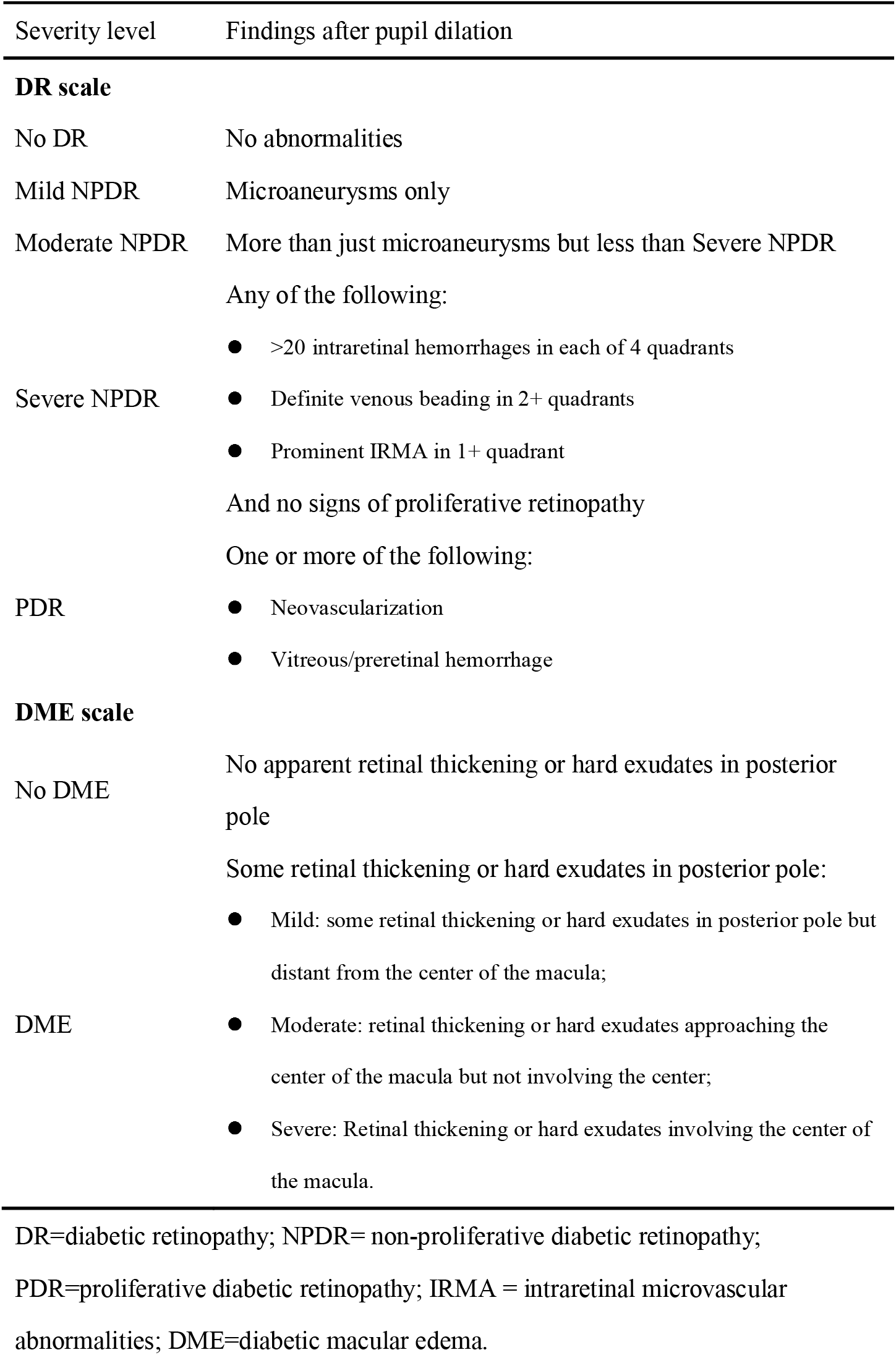
International clinical disease severity scale for diabetic retinopathy (DR) and diabetic macular edema (DME).

Optical coherence tomography (OCT): The retina was examined with Heidelberg Spectralis (Heidelberg Engineering, Heidelberg, Germany) to perform cross-sectional scanning of the macular retina using a 9 mm horizontal line and a 6mm×6mm volume scan model. The optic nerve head was imaged with a 3.4 mm circle scan mode and a 6 mm radical scan mode. A commercial swept source OCTA device (Triton DRI-OCT, Topcon. Inc., Tokyo, Japan) was used by experienced and skilled doctors to image the blood flow, with a wavelength of 1050 nm within a range of 100 nm and an achievable horizontal resolution of 8 μm. Blood flow was visualised using the 3×3mm model, which was automatically stratified into the superficial, deep, outer and choroid-capillary layers.

### Questionnaire interview

Detailed questionnaire interviews were administered by surveyors to collect data on personal information, lifestyle risk factors and disease history. Personal information included the date of birth, education background, career and income. Lifestyle risk factors included dietary habits, smoking and consumption of alcohol. A condensed and translated version of the International Physical Activity Questionnaire (IPAQ) was used to assess physical activity, and the Pittsburgh Sleep Quality Index (PSQI) was employed to evaluate sleep quality.^17,18^ Disease history included the participants’ systemic diseases, long-term use of medication, ocular diseases and surgeries.

### Biological sample collection

All the participants who agreed to participate in the study were instructed to fast for at least 8 hours before blood sampling. A trained nurse collected overnight fasting venous blood samples to test for HbA1c, creatinine, uric acid and lipids (total cholesterol, triglycerides, high-density and low-density lipoprotein cholesterol). The whole blood sample with the EDTA anticoagulant was centrifuged at 3000 rpm for 10 minutes to obtain plasma. During processing, all the samples were kept from exposure to direct light. Genomic DNA was also collected from the blood samples with the EDTA anticoagulant. The extracted samples were stored in vacuum tubes and kept at −80°C for possible later molecular studies. The urine samples were collected for a quantitative measurement of urinary albumin.

Physical parameters and blood pressure measurement: The parameters included weight, height and waist and hip circumference. Weight and height were measured with an automatic weight and height scale, with the participants standing on the scale with light clothes and no shoes on. The body mass index (BMI) was defined as the weight (kg) divided by the square of height (m). The waist-height ratio (WHR) was calculated as the hip circumference divide by the waist circumference. Systolic and diastolic blood pressure values were measured by a blood pressure monitor three times at 5-minute intervals with participants in quiescent conditions.

### Data management and quality control

Quality control was implemented throughout the GDES to maximise the accuracy and completeness of the final database. All staff members received comprehensive and detailed training and only those with proven competence participated into the study. It was ensured that all staff members mastered a good knowledge of the study purpose, procedures, data collection forms, and skill to perform examinations to secure compliance to and the scientificity of the study.

The measures for data management and quality control included the following: (1) Standardized case-report form (CRF) was adopted to ensure all the examinations were completed and certain data (such as refraction, visual acuity and IOP) were recorded; (2) the completeness of the questionnaire interview was checked before the participants left ZOC; if the CRF was not completely filled in, the staff in charge of the examinations was asked to fill in the missing results or resolve any noticed data discrepancy; (3) all the examination results were synchronised and backed up to an independent internet-based server; (4) all the data from the paper-based CRFs were entered into an electronic database after the completion of the baseline visit; and (5) all the paper-based CRFs were stored in locked cabinets to secure the privacy of the participants.

Measures to improve compliance for the follow-up visits included the following: (1) The participants included in the study were examined free of charge, and coupons for spectacle dispensing were gifted; (2) as much contact information of the participants as possible was obtained, including the home address, the workplace address, land line numbers, mobile phone numbers, and other methods through which he/she could be contacted (as well as contact information of the family members); and (3) follow-up reminders were sent via messages to the participants prior to the next follow-up visit.

All the subjective clinical examinations were performed by experienced ophthalmologists and designated technicians. All the graders were strictly trained and tested for repeatability and reproducibility of images grading (kappa value ≥ 0.9).^9^ All the examinations were carried out in accordance with the standard operating procedures (SOP) developed prior to the study. A project leader randomly checked the data of 10 participants on a monthly basis to identify and solve any possible problems in a timely manner. All the equipment and devices were regularly calibrated according to the instructions and manuals to ensure accuracy and consistency.

### Statistical analyses

The eye in the worst condition was included in the analysis. Data processing analysis was performed using STATA statistical software (Stata version 12.0, Stata Corp., College Station, TX). The categorical variables were presented as numerical percentages; the normal distribution of the continuous variables was presented using mean ± standard deviation (SD), and the non-positive distribution of continuous variables was expressed as median (interquartile range) values. Normal distribution was confirmed using the Kolmogorov-Smirnov test. The associations between demographic characteristics or lifestyle factors at baseline and presence of DR were explored using a logistic regression analysis to identify possible risk factors. The following factors were used as independent variables for stepwise model, including age, sex, duration of DM, HbA1c, use of insulin, BMI, MBP, total cholesterol, serum creatinine, HDL-c, LDL-c, triglycerides, serum uric acid, C-reactive protein, MAU, current smoker, alcohol intake, sleep duration, and metabolic equivalents. P-values, odds ratios (ORs) and 95% confidence intervals (CIs) were used to show the statistical results. A P-value < 0.05 was considered statistically significant.

## Results

### Demographic characteristics of the study population

A total of 2716 potentially eligible patients with diabetes mellitus in the community health centres were invited, and 2564 (94.4%) participants arrived at the Clinical Research Centre in Zhongshan Ophthalmic Centre at the end of the baseline (Dec 30, 2019). Figure 1 shows the process of GDES. Among of the 2511 (participate rate 92.5%) patients completed the questionnaires and examinations, 2372 (87.3%) were eligible for inclusion. Retinal photographs of 67 participants were ungradable. Therefore, this study finally included 2305 (84.9%) patients in the final analysis. During the process of quality control, only one patient’s data was found to be wrong, and we made timely corrections. There were 990 (42.95%) male, and 1315 (57.05%) female, with a mean age of 64.4±7.8 years, a duration of DM of 8.8±7.0 years and a mean HbA1c concentration of 7.04%±1.44%. The numbers of participants with a history of hypertension, hyperlipidemia and articular gout were 1309 (56.79%), 787 (34.14%) and 231 (10.02%), respectively.

**Figure 1.**
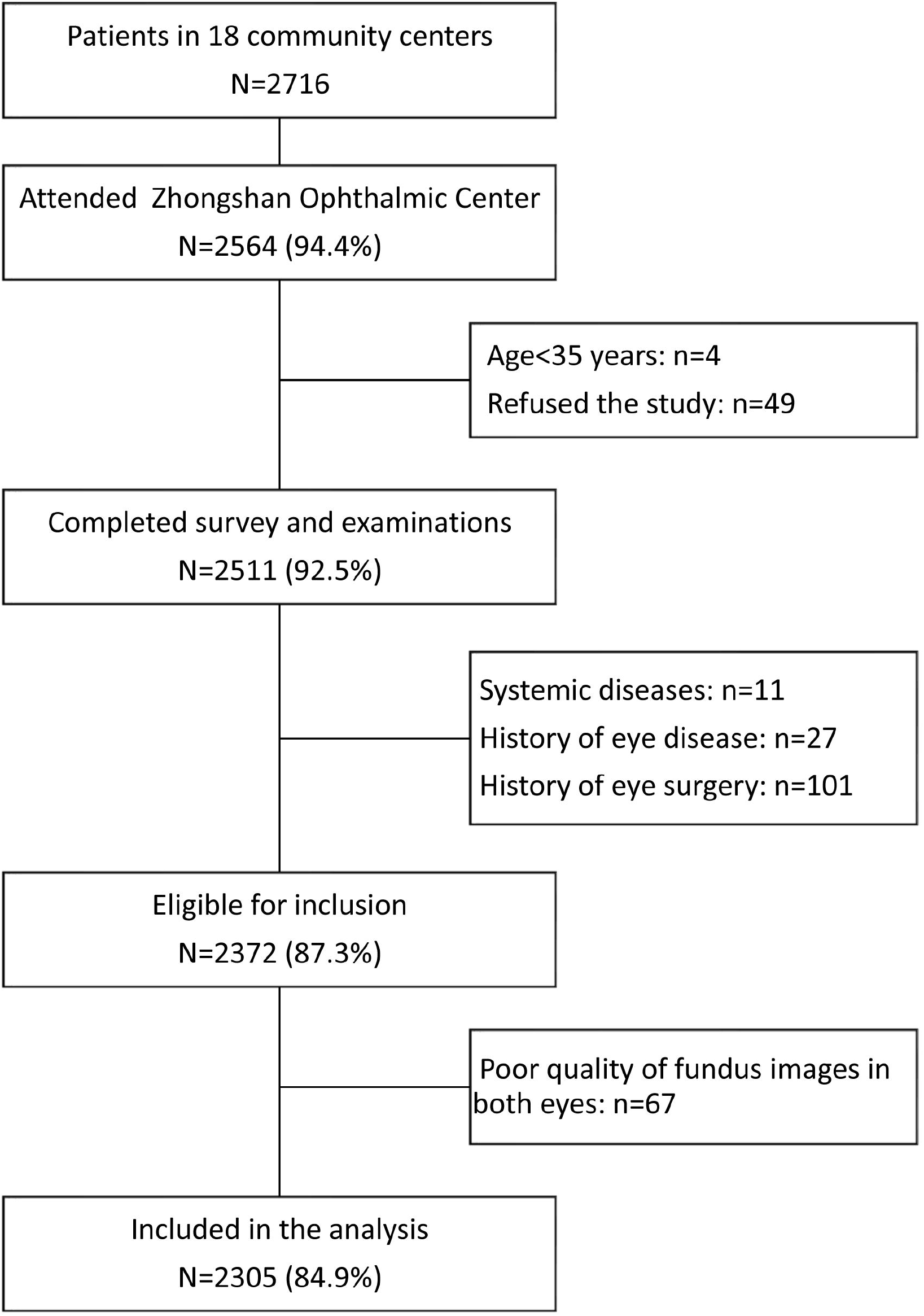
Flowchart of the Guangzhou Diabetic Eye Study.

### Baseline characteristics of DR according to DM duration

Table 21 shows the prevalences of DR according to the durations of DM. In total, 14.58% of the participants had any DR and 4.25% had VTDR, among which 76 (3.30%), 197 (8.55%), 45 (1.95%) and 17 (0.74%) were classified as mild NPDR, moderate NPDR, severe NPDR and PDR, respectively. There were 93 (4.03%) patients of DME. When the participants were stratified by duration of DM, the prevalence of any DR and VTDR among diabetic patients with a duration of less than 5 years was 7.32% and 2.40%, respectively, and 33.18% and 7.94% among those with more than 20 years.

### Independent risk factors for any DR and VTDR

Table 3 presents the results of the multiple logistic regression analysis to reveal the independent risk factors of DR and VTDR, the former of which included a longer duration of DM (OR = 1.59, 95% CI: 1.38–1.83, P < 0.001), higher degree of HbA1C (OR = 1.45, 95% CI: 1.34–1.56, P < 0.001), insulin treatment (OR = 2.31, 95% CI: 1.75–3.05, P < 0.001), higher average arterial pressure (OR = 1.01, 95% CI: 1.004–1.02, P = 0.001), higher concentration of serum creatinine (OR = 1.02, 95% CI: 1.01–1.02, P < 0.001) and the presence of urinary microalbumin (OR = 1.004, 95% CI: 1.001–1.007, P = 0.010). The independent protection factors of DR included younger age (OR = 0.96, 95% CI: 0.95–0.98, P <0.001) and higher BMI (OR = 0.95, 95% CI: 0.91–0.99, P = 0.022).

**Table 2.** Prevalence of DR according to duration of diabetes mellitus.

| DR grading | DM duration (years) |  |  |  | Total |
| --- | --- | --- | --- | --- | --- |
|  | < 5 | 5-10 | 10-20 | ≥ 20 |  |
| NDR | 772 (92.68%) | 515 (88.34%) | 539 (79.85%) | 143 (66.82%) | 1970 (85.42%) |
| Any DR | 61 (7.32%) | 68 (11.66%) | 136 (20.15%) | 71 (33.18%) | 336 (14.58%) |
| Mild NPDR | 19 (2.28%) | 15 (2.57%) | 30 (4.44%) | 12 (5.61%) | 76 (3.30%) |
| Moderate NPDR | 29 (3.48%) | 46 (7.89%) | 79 (11.7%) | 43 (20.09%) | 197 (8.55%) |
| Severe NPDR | 9 (1.08%) | 5 (0.86%) | 22 (3.26%) | 9 (4.21%) | 45 (1.95%) |
| PDR | 4 (0.48%) | 2 (0.34%) | 4 (0.59%) | 7 (3.27%) | 17 (0.74%) |
| DME | 20 (2.40%) | 16 (2.74%) | 42 (6.22%) | 15 (7.01%) | 93 (4.03%) |
| VTDR | 20 (2.40%) | 17 (2.92%) | 44 (6.52%) | 17 (7.94%) | 98 (4.25%) |

2 DM=diabetes mellitus; DR=diabetic retinopathy; NDR=non diabetic retinopathy; NPDR=non proliferative diabetic retinopathy; VTDR=vision 3 threatening diabetic retinopathy; DME=diabetic macular edema.

**Table 3.** Stepwise multivariable logistic regression analyses of independent factors associated with any DR, VTDR, and DME.

| Factors* | Any DR |  | VTDR |  | DME |  |
| --- | --- | --- | --- | --- | --- | --- |
|  | OR (95%CI) | P-value | OR (95%CI) | P-value | OR (95%CI) | P-value |
| Age, year | 0.96 (0.95, 0.98) | <0.001 | 0.94 (0.92, 0.97) | <0.001 | 0.93 (0.91, 0.96) | <0.001 |
| Duration of DM, per 5-year | 1.59 (1.38, 1.83) | <0.001 | 1.46 (1.15, 1.85) | 0.002 | 1.43 (1.12, 1.83) | 0.004 |
| HbA1c, % | 1.45 (1.34, 1.56) | <0.001 | 1.38 (1.23, 1.55) | <0.001 | 1.35 (1.20, 1.52) | <0.001 |
| Use of insulin, yes vs no | 2.31 (1.75, 3.05) | <0.001 | 2.01 (1.27, 3.18) | 0.003 | 1.95 (1.21, 3.12) | 0.006 |
| BMI, kg/m<sup>2</sup> | 0.95 (0.91, 0.99) | 0.022 | 0.90 (0.84, 0.97) | 0.004 | 0.90 (0.84, 0.96) | 0.003 |
| MBP, mmHg | 1.01 (1.004, 1.02) | 0.001 | – | – | 1.011 (1.002, 1.020) | 0.020 |
| Serum creatine, µmol/L | 1.02 (1.01, 1.02) | <0.001 | 1.01 (1.003, 1.02) | 0.006 | 1.010 (1.003, 1.017) | 0.007 |
| Microalbuminuria, mg/mL | 1.004 (1.001, 1.007) | 0.010 | 1.005 (1.002, 1.008) | 0.001 | 1.005 (1.002, 1.008) | 0.003 |

7 DM=diabetes mellitus; BMI=body mass index; MBP=mean blood pressure; OR=odds ratio; 95%CI=95% confidential interval.
\*Age, sex, duration of DM, HbA1c, use of insulin, BMI, MBP, total cholesterol, serum creatinine, HDL-c, LDL-c, triglycerides, serum uric acid,
9 C-reactive protein, MAU, current smoker, alcohol intake, sleep duration, and metabolic equivalents were used for stepwise multivariable model.

For VTDR, the following seven factors were of vital importance, including older age (OR = 0.94, 95% CI: 0.92–0.97, P < 0.001), a longer duration of DM (OR = 1.46, 95% CI: 1.15–1.85, P = 0.002), higher concentration of HbA1c (OR = 1.38, 95% CI: 1.23–1.55, P < 0.001), use of insulin (OR = 2.01, 95% CI: 1.27–3.18, P = 0.003), lower BMI (OR = 0.90, 95% CI: 0.84–0.97, P = 0.004), higher concentration of serum creatinine (OR = 1.01, 95% CI: 1.003–1.02, P = 0.006) and higher urinary microalbumin (OR = 1.005, 95% CI: 1.002–1.008, P = 0.001). Stepwise multiple logistic regression demonstrated that age, duration, use of insulin, BMI, MBP, serum creatine, MAU were also independent factors for DME (Table 2). Linear regression analyses showed that the BMI was negatively correlated with duration of disease (β = −0.049; 95% CI: −0.068, −0.029; P <0.001), and showed no correlation with HbA1c (β = 0.076; 95% CI: −0.017, 0.170; P = 0.109) and use of insulin (β = −0.102; 95% CI: −0.429, 0.225; P = 0.541).

## Discussion

The GDES is a community-based cohort study in which samples representative of the diabetic population living in southern urban China were collected. The main purpose of this study is to better understand the onset and natural progression of DR in type 2 diabetic population in southern urban China to reveal the relevant environmental and genetic risk factors and to establish a DR prediction model based on the identified risk factors. The relatively large sample size of the community-based type 2 diabetic cohort was successfully recruited, and the subsequent follow-up visits will provide insights on the early prevention of DR for individuals at high risk.

The findings of our study are consistent with those reported in the previous epidemiology studies.^2, 19-22^ For instance, the Singapore Eye Study reported similar DR risk factors despite the ethnic variations in DR prevalence. The main risk factors for DR included a longer duration of DM, younger age of DM diagnosis, higher concentration of HbA1c, higher blood pressure, the use of insulin and the presence of albuminuria.^2^ Correlations among cases of new DR onset with higher concentrations of HbA1c, longer duration of DM, higher serum creatinine concentrations and so forth were observed in the Beijing Eye Study, which was consistent with a meta-analysis of cross-sectional studies of the Chinese population reporting younger age at diagnosis, longer duration of disease, the use of insulin and fasting blood sugar and HbA1c levels as independent risk factors.^23, 24^ A controversy has existed regarding the association between BMI and DR; the study in Singapore reported BMI as a protection factor of DR, whereas a risk factor identified in studies in Japan and northern China and bearing no association at all in other studies.^25-27^ We analysed the relationship between BMI and duration of disease, insulin use, and HbA1c concentration and found that BMI was negatively correlated with duration of disease. This result indicated that a shorter exposure to hyperglycemia may contribute to the protective effect of BMI on DR. The findings of our study are similar to those in the Singapore study, but longitudinal data are warranted to resolve the dispute.

With only one population-based study, two community-based studies and one hospital-based study, cohort studies of DR in the Chinese diabetic population are rare.^6, 7, 28, 29^ The Beijing Eye Study is a population-based study reported that the 10-year incidence of any DR was 4.2% among 246 diabetic patients in northern China.^28^ The prospective research in the Xinjing Community in Shanghai enrolled 778 participants with type 2 diabetes, and reported that the 5-year incidence of DR was 46.89%.^6^ The researchers in another community-based cohort study conducted in Shenyang, Liaoning, recruited 212 type 2 diabetic patients and observed an 5-year incidence of DR of 9.05%.^7^ A hospital-based cohort study of diabetic patients in Hong Kong included 5160 patients. Among the 3647 patients without DR at baseline, the 4-year incidence of any DR, mild NPDR, moderate NPDR and VTDR were 15.16%, 14.45%, 0.69% and 0.03%, respectively. Among the 1513 patients with DR at baseline, the 4-year progression rate of DR was 6.61% and the regression rate was 45.54%.^29^ Apparent discrepancies prevail among these studies, which might be attributed to the variations of demographic characteristics, lifestyles, healthcare and economic well-being of the study samples. All the above studies lacked ETDRS-7-field retinal photographs and genotyping, which were all collected in the GDES, and the subsequent follow-up study will enrich the information on the mechanisms underlying DR onset and progression in the Chinese population.

Recently, new ideas have been raised in the pathology of DR. The geometric distribution of blood flow and blood vessels in the macula and the optic papilla can be quantified by the newly-developed OCTA device, thus opening up new fields for future research.^30^ In some studies, it has been reported that changes of blood flow preceded the development of DR symptoms in diabetic patients, contrary to some other studies.^31^ As most of the data were extracted from studies of limited sample size, and few have reported on the association between OCTA parameters and the genetic risk factors of DR, the new technologies applied in our study (e.g. OCTA and SNP) were able to bridge such gaps.

The strengths of our studies included the following: (1) The participants were recruited from a single district of relative high stability, thus cutting down the confounding influence of environmental factors and the attrition rate; (2) direct comparisons with studies of Western populations are possible with the application of the ETDRS-7-field retinal photographs and the grading scheme, which in turn facilitate academic exchange; (3) the number of participants was far larger than the needed sample size, and the results offer insights into the onset and progression of DR in the Chinese population; and (4) the establishment of blood plasma and molecular database might accelerate the investigation of genetic and non-genetic factors related to the onset and progression of DR. However, some shortcomings should be noted: (1) The inclusion of participants was not population-based sampling, which might give rise to selection bias, and thus the association between risk factors and DR cannot be generalised to the entire population. However, we enrolled diabetic patients registered in community service centres in Yuexiu District, Guangzhou. All the 18 community centers were included with no random sampling. (2) a fluorescein angiography examination was lacking, as it was not feasible to perform it in such a large-scale population; (3) the course of diabetes may be underestimated, because of a large number of postponed diagnostics of diabetic patients in China. Finally, the duration of diabetes may be underestimated, because of a large number of postponed diagnostics of diabetic patients in China. The International Diabetes Federation (IDF) estimated that 56.0% of diabetic patients in China have not been diagnosed in time.^32^ In the Yangxi Eye study, we found that 85.9% of diabetic patients were not diagnosed in rural area, which led to a shortened calculation of the duration of diabetes.^9^

In summary, this study described the methodology and characteristics of GDES, which is an ongoing prospective cohort study of the type 2 diabetic population in southern urban China. The same standardised grading scheme as that applied in studies of Western populations was adopted, as well as the newly-developed ocular imaging modality. The results at baseline showed that DR prevalence and the major risk factors in the Chinese population were consistent with those in studies with Western populations. The results can be used to better inform policy-making on DR prevention and treatment.

## Data Availability

Available when appropriate request.

## Acknowledgements

We thank all participants and related staffs of this study.

## Author Contributions

WW and WH had full access to all the data in the study and take responsibility for the integrity of the data and the accuracy of the data analysis. Study concept and design: WW, WL, MH, WH. Acquisition, analysis, or interpretation of data: XG, LW, JM, YL, KX, WL. Drafting of the manuscript: WW, MH. Critical revision of the manuscript for important intellectual content: All authors. Statistical analysis: WW. Obtained funding: WH. Administrative, technical, or material support: MH, WW. Study supervision: MH.

## Data Availability

All relevant data are within the paper.

## Patient consent

Obtained.

## Ethics approval

This study was approved by the Ethics Committee of Zhongshan Ophthalmic Center, Sun Yat-sen University, Guangzhou, China.

